# Impact of spatiotemporal heterogeneity in COVID-19 disease surveillance on epidemiological parameters and case growth rates

**DOI:** 10.1101/2022.03.31.22273230

**Authors:** Rhys P.D. Inward, Felix Jackson, Abhishek Dasgupta, Graham Lee, Anya Lindström Battle, Global.health consortium, Kris V. Parag, Moritz U.G. Kraemer

## Abstract

SARS-CoV-2 case data are primary sources for estimating epidemiological parameters and for modelling the dynamics of outbreaks. Understanding biases within case based data sources used in epidemiological analyses are important as they can detract from the value of these rich datasets. This raises questions of how variations in surveillance can affect the estimation of epidemiological parameters such as the case growth rates. We use standardised line list data of COVID-19 from Argentina, Brazil, Mexico and Colombia to estimate delay distributions of symptom-onset-to-confirmation, -hospitalisation and -death as well as hospitalisation-to-death at high spatial resolutions and throughout time. Using these estimates, we model the biases introduced by the delay from symptom-onset-to-confirmation on national and state level case growth rates (rt) using an adaptation of the Richardson-Lucy deconvolution algorithm. We find significant heterogeneities in the estimation of delay distributions through time and space with delay difference of up to 19 days between epochs at the state level. Further, we find that by changing the spatial scale, estimates of case growth rate can vary by up to 0.13 d^-1^. Lastly, we find that states with a high variance and/or mean delay in symptom-onset-to-diagnosis also have the largest difference between the rt estimated from raw and deconvolved case counts at the state level. We highlight the importance of high-resolution case based data in understanding biases in disease reporting and how these biases can be avoided by adjusting case numbers based on empirical delay distributions. Code and openly accessible data to reproduce analyses presented here are available.

## Introduction

Surveillance of Severe acute respiratory syndrome coronavirus 2 (SARS-CoV-2) has expanded since it was first reported in November 2019 (Oude Munnink *et al*., 2021; Zhu *et al*., 2020). However, disease surveillance remains highly heterogeneous across countries and case definitions have changed significantly as a result of changing testing capacity, improved understanding about transmission during the asymptomatic phase and general human behavioural change in response to the pandemic (Flaxman *et al*., 2020; Verity *et al*., 2020; Wu *et al*., 2020; Ke *et al*., 2021; Pullano *et al*., 2021; Parag, Cowling and Donnelly, 2022). Improvements to surveillance efforts can affect key epidemiological distributions by reducing the time delay from exposure to onset of infectiousness to diagnosis (Kraemer *et al*., 2021). These in turn can directly influence estimation of the time-varying reproduction number (R_t_) and growth rate (r_t_) (Rong et al., 2020; Pitzer et al., 2021) (Supplementary Table. 1). Estimation of these epidemiological distributions/parameters provide key information on changes in transmission, which contribute to decisions on the implementation of pharmaceutical and non-pharmaceutical interventions (NPIs) (Anderson *et al*., 2020; Dushoff and Park, 2021; Parag, Thompson and Donnelly, 2021; Pellis *et al*., 2021).

Initial estimations of SARS-CoV-2 epidemiological distributions/parameters were based on biased data primarily due to limited capacity of testing for SARS-CoV-2 in hospitalised patients (Vandenberg *et al*., 2021). This contributes to a degree of uncertainty and heterogeneity in the accuracy and precision of these estimates especially when comparing them between countries and across age groups (Cowling et al., 2020; Mellan et al., 2020; Verity et al., 2020; Parag, Cowling and Donnelly, 2022). Since the initial stages of the pandemic, global surveillance and notification systems have significantly improved (Vandenberg *et al*., 2021) providing a wealth of data which can be used to re-evaluate SARS-CoV-2 epidemiological distributions/parameters.

This raises the question of how variations in surveillance affects the estimation of epidemiological distributions/parameters. We aim to understand how spatial and temporal heterogeneities in reporting (specifically delays in reporting) can impact the accuracy of estimates of epidemiological parameters (specifically growth rate *r*_*t*_) within and between countries. To do this, we are using a rich, standardised, and individual level line list database extracted from Global.health (https://global.health/). We focus on estimating the delays between symptom-onset-to-confirmation, -hospitalisation and -death as well as hospitalisation-to-death.

## Methods

### Data

The Global.health database contains individual case data from over 100 countries (https://global.health/). The database contains a rich array of fields describing demographics, location (up to Administrative Area 3 resolution), and key epidemiological and clinical events for confirmed COVID-19 cases. In relational database format, each row is a single confirmed COVID-19 case, and columns detail attributes for each case (Schema: https://github.com/globaldothealth/list/blob/c0da57d6b227ab861ad5e695d711699c02c2721f/data-serving/scripts/export-data/data_dictionary.txt). Data is primarily sourced from official country line lists compiled and shared by national health institutions where available, as was the case for all countries in this study (Xu *et al*., 2020). The detail of the case data varies by country: inter-country variability in COVID-19 data collection and reporting online leads to differences in Global.health data availability, as detailed in Figure 1. The dataset used in this study was downloaded from Global.health on 31/01/2022. An updated line list can be downloaded from Global.health via the website or by following instructions on the API docs: https://github.com/globaldothealth/list/tree/main/api. We can provide the exact dataset downloaded for this analysis upon written request.

**Figure 1:**
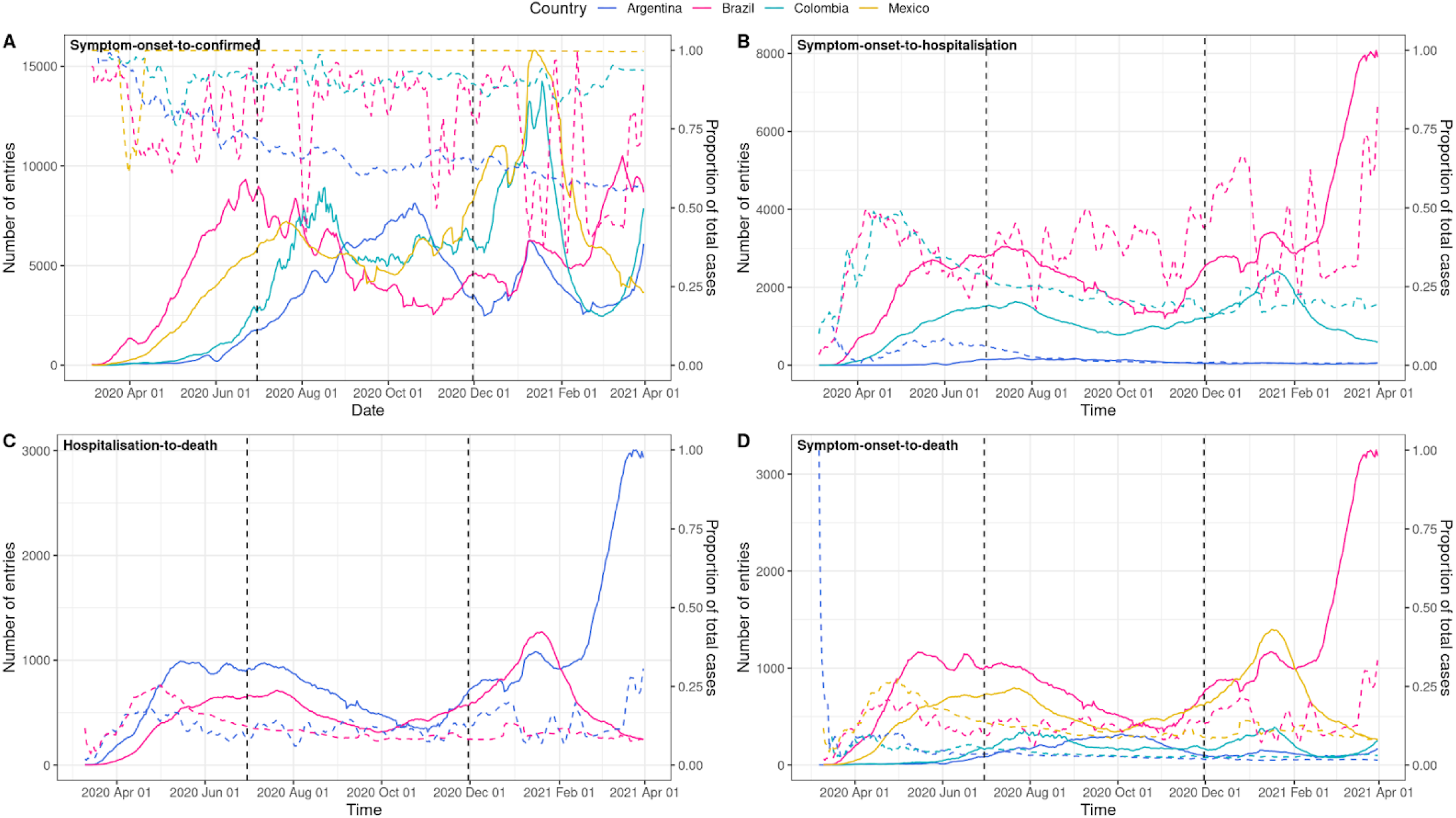
The number and proportion of recorded cases with data entries for each epidemiological distribution have been extracted from Global.health line lists for Argentina, Brazil, Colombia, and Mexico. Figure 1A, 1B and 1D represent the delay from symptom-onset-to-diagnosis, -hospitalisation, and -death respectively whilst Figure 1C represents the delay from hospitalisation-to-death. The blue, red, teal, and yellow solid line represents a 7-day rolling average for the total number of data entries for Argentina, Brazil, Colombia, and Mexico respectively. The blue, red, teal and yellow dashed line represents a 7 day rolling average for the proportion of recorded cases with data entries for Argentina, Brazil, Colombia, and Mexico respectively. The dashed vertical lines represent epoch change times.

To investigate the spatial heterogeneity of epidemiological parameters inferred from public data, we focus on COVID-19 line lists from four countries in Latin America that have consistently provided comprehensive and detailed line list data since the start of the pandemic in early 2020: Mexico, Brazil, Argentina, and Colombia. For each country, we aggregated data to the state level, then for each state, calculated delay distributions defined in Supplementary Table 1. To investigate trends over time, the line lists for each country are split into three time-periods hereafter called epochs. These epochs represent different stages of the SARS-CoV-2 epidemic in each country. We cover the 1st and 2nd waves of infections as well as a period of low incidence in infections between these two waves:

- **Epoch 1:** 2020-03-03 to 2020-06-30 (initial COVID-19 wave)
- **Epoch 2:** 2020-07-01 to 2020-11-30 (receding epidemic and low case counts)
- **Epoch 3:** 2020-12-01 to 2021-03-31 (second wave/SARS-CoV-2 VOCs)

Additional filtering of the data was applied to these time delays to eliminate biases introduced by erroneous entries. We removed all cases which were reported before the first reported case in the countries of interest based on the Ministry of Health’s websites (Roberts, Rossman and Jarić, 2021). Moreover, we removed outliers that fell outside of the 97.5% range of the data on each of the delay distributions.

### Epidemiological Distributions

To estimate the epidemiological distribution, a gamma probability density function (PDF) was fitted to onset-to-death and hospitalisation-to-death whilst a generalised lognormal (GLN) probability density function (Singh *et al*., 2012) was fitted to onset to diagnosis and hospitalisation (Table 1). These PDFs were chosen as they were evaluated to best fit COVID-19 line list data (Hawryluk *et. al*., 2020). The parameters of each distribution are fitted by a joint hierarchical model with partial pooling similar to (Hawryluk *et. al*., 2020), using state level data (Administrative Area 1 resolution) from Argentina, Brazil, Colombia, and Mexico.

**Table 1:**
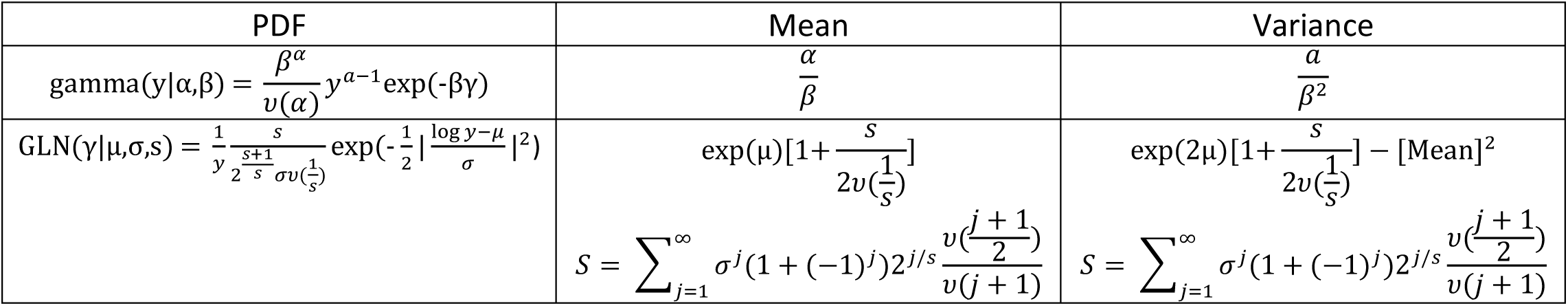
Probability density functions with analytical formulae for mean and variance. y denotes the data, ***v*** () is a gamma function. GLN, generalised log-normal.

Posterior samples of the parameters are generated using Hamiltonian Monte Carlo (HMC) (Hoffman and Gelman, 2014) in Stan (Carpenter *et al*., 2017) using PyStan (v.2.19.0.0: https://mc-stan.org/users/interfaces/pystan). Four chains with 2000 iterations, with 50% of the iterations dedicated to burn-in, were used for each fit. For all fitted densities, the mean and variance parameters were constrained to be positive.

### Correlation analysis

Spearman’s rank-order correlation coefficient (r_s_) was calculated for delays between symptom-onset-to-confirmation, -hospitalisation and -death as well as hospitalisation-to-death for each state, using the scipy.stats ‘spearmanr’ function (scipy version 1.7.3). P-values are provided by this function, which indicates the probability of an uncorrelated system producing data with a correlation value at least as extreme as the one observed. The p-values should be interpreted with caution as we have a limited sample size (n = number of states in each country).

### Deconvolution

We used deconvolution to adjust for delays in the development of detectable viral loads, symptom onset, and reporting (Gostic *et al*., 2020). Deconvolution allows us to reconstruct the unlagged incidence time series given a known delay distribution (estimated above). Here, we adapted the method by Goldstein et al. (Goldstein *et al*., 2009). This method uses the daily confirmed incidence curve (I_t_) and the symptom onset to confirmation probability distribution (d_1,…,_d_I_) to calculate the expected number of cases (μ_t_) to occur at time *t* adjusting for delays. We assume that the daily incidence curve (I_t_) is Poisson distributed. The model requires non-negativity constraints on the parameters *λ*_*t*_, which represents estimates of mean infection incidence, reflecting the fact that they are Poisson means.

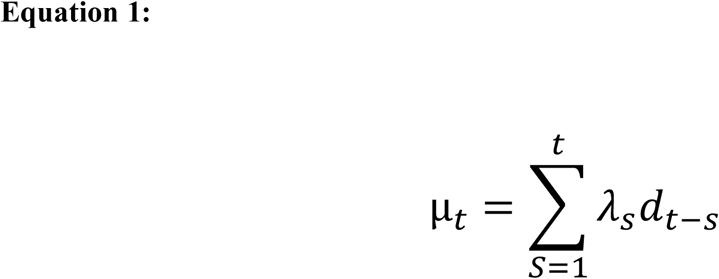

The model ran for 50 iterations or until the normalised x^2^ statistic (Equation 2) comparing the observed and expected number of cases per day falls below 1. Here, N represents the length of our study period, E is the expected number of cases on day i and D is the probability of observation on day i. We calculated the deconvolved case counts at both the national and state level for each epoch.

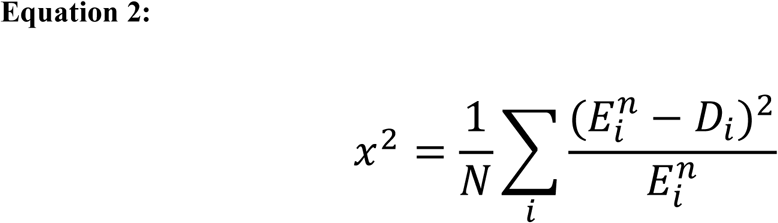

### Growth rate

To estimate the daily growth rate (*r*_*t*_) by country and state we adapted the approach from Pellis *et al*. (Pellis *et al*., 2021). In short, the growth of daily case numbers of lagged and unlagged SARS-CoV-2 cases (*y*) at time (*t*) was considered exponential. To estimate *r*_*t*_, a Poisson family generalised linear model (GLM) with a log link was applied. To allow growth rates to vary over time in a semi-parametric manner, a generalised additive model (GAM) was used where y(*t*) ∝ e^s(t)^ for some smoother s(*t*). As such, *r*_*t*_ is the time derivative of the smoother *r*_*t*_ = s(*t*). We started calculating the growth rate once the cumulative number of daily cases reached over 100 on the national level and over 20 on the state level to ensure that the exponential growth phase was captured.

Code: Code to reproduce analyses can be accessed here:

https://github.com/fojackson8/COVID19_mapping_epiparams and data can be downloaded via https://data.covid-19.global.health/ or via our API: https://github.com/globaldothealth/list/tree/main/api. Data downloads require agreeing with the Terms of Use: https://global.health/terms-of-use/.

## Results

### Number of Data Entries / Global.health Case Counts

Disease reporting varied by country and field. Figure 1 shows the number and proportion of recorded cases with data entries from the Global.health linelist from which we can infer the delays between onset-to-confirmation (A), onset-to-hospitalisation (B), hospitalisation-to-death (C) and onset-to-death (D). There are significant heterogeneities between countries and overtime between the number of cases recorded and a data entry being present for a specific delay. For example, almost all cases in Mexico are populated with the delay between onset-to-confirmation. In contrast, while almost all initial cases in Argentina were populated with the delay between onset-to-confirmation, over time, the proportion of cases with data entries fell consistently to around 55%. Further, there is a large variability in completeness of the fields that allow estimation of symptom-onset-to-diagnosis ranging between 36% - 97% in Brazil.

### Estimation of Delay distribution and Growth rate

We estimate the delay distributions (Supplementary table 1), reconstruct deconvolved case numbers and *r*_*t*_ for local SARS-CoV-2 epidemics in Argentina, Brazil, Colombia, and Mexico.

### Delay Distributions

PDFs were applied to epidemiological data from Argentina, Brazil, Colombia, and Mexico to estimate the delay from symptom onset-to-diagnosis, delay from symptom onset-to-hospitalisation, delay from hospitalisation-to-death, and the delay from symptom onset-to-death at the state level. Posterior plots of state-level results (Figures 2-3 and Supplementary Figures 2-3) show the shape and spread for the delay for all delay distributions between states and over time.

**Figure 2:**
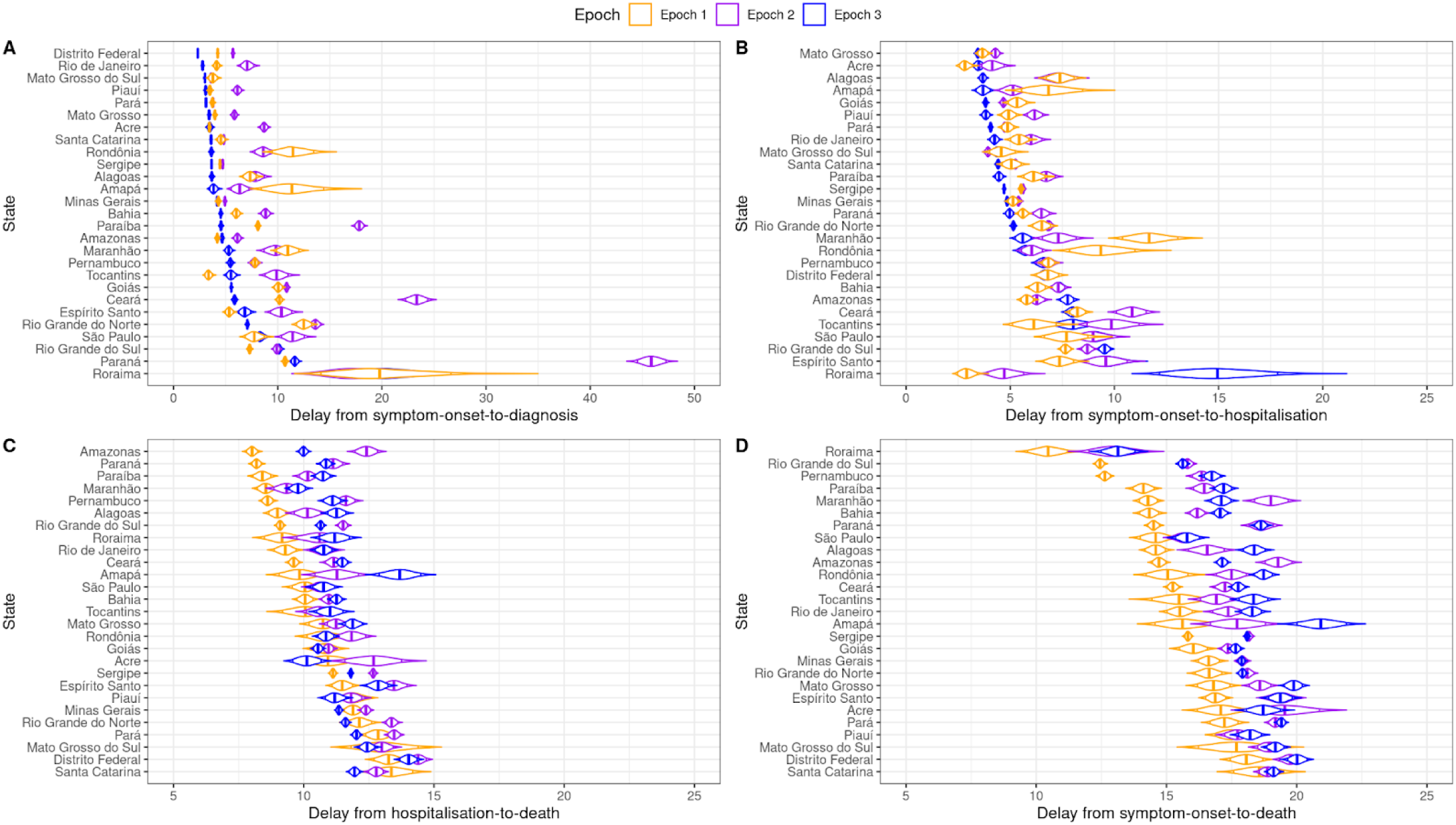
Delay distributions are estimated from daily case counts on the state level for three distinct epochs for Brazil. Figure 2A, 2B and 2D represent the delay from symptom-onset-to-diagnosis, -hospitalisation, and -death respectively whilst Figure 2C represents the delay from hospitalisation-to-death. Orange represents epoch 1, purple represents epoch 2 and blue represents epoch three. All plots are ordered from the smallest to largest by the epoch with the smallest mean delay.

**Figure 3:**
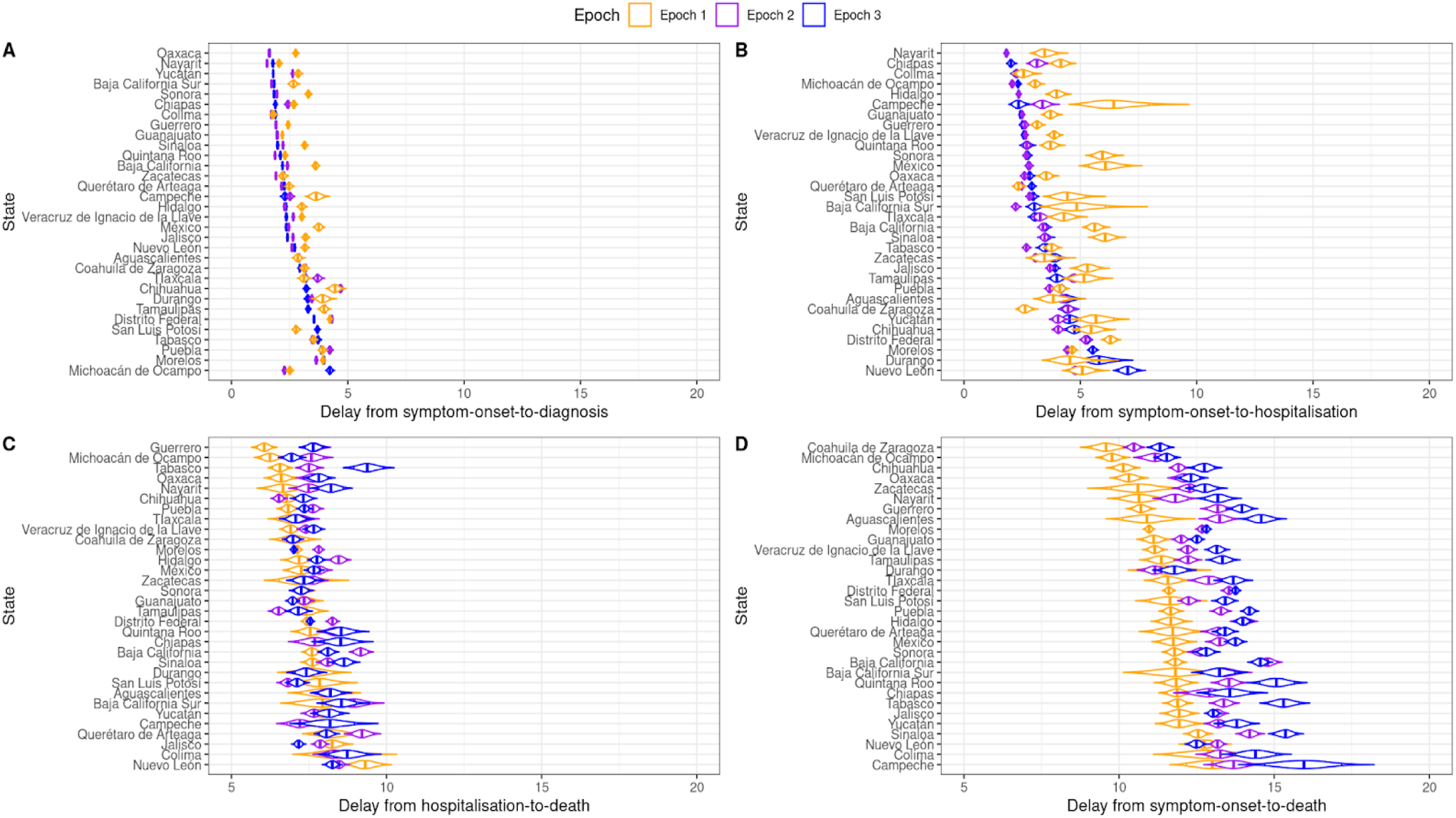
Delay distributions are estimated from daily case counts on the state level for three distinct epochs for Mexico. Figure 3A, 3B and 3D represent the delay from symptom-onset-to-diagnosis, -hospitalisation, and -death respectively whilst Figure 3C represents the delay from hospitalisation-to-death. Orange represents epoch 1, purple represents epoch 2 and blue represents epoch three. All plots are ordered from the smallest to largest by the epoch with the smallest mean delay.

#### Brazil

In Brazil, we observe substantial heterogeneities in the mean delay across all four distributions between states and for the epochs. For example, for all states, the mean delay from symptom-onset-to-diagnosis increases from 7.24 days in epoch 1 to 10.46 days in epoch 2, declining to 5.55 days in epoch 3 (Supplementary Table 2). At the state level, Distrito Federal had the 3rd overall lowest mean delay of 4.08 days whilst Paraná had the highest mean delay of 22.74 days (Figure 2, Supplementary Table 3). Interestingly, this trend was reversed for the distribution of hospitalisation-to-death with Distrito Federal having the highest mean delay of 13.89 days and Paraná having the 3rd lowest mean delay of 10.01 days (Figure 2, Supplementary Table 3). Additionally, states with a large delay from symptom-onset-to-diagnosis also had a large delay from symptom-onset-to-hospitalisation (r_s_ = 0.58, p < 0.01). Conversely, we found states with a large delay from symptom-onset-to-diagnosis had a shorter delay from hospitalisation-to-death (r_s_ = 0.60, p < 0.01) (Supplementary Figure 1). Moreover, we found that the longer the delay from symptom-onset-to-hospitalisation the shorter the delay from hospitalisation-to-death (r_s_ = -0.37, p < 0.01) (Supplementary Figure 1) implying the longer it takes to be hospitalised after becoming symptomatic the shorter the time in hospital before death.

#### Mexico

Similar to Brazil, we found heterogeneities across states and time for all delay distributions within Mexico (Figure 3). Moreover, the trends for each distribution overtime are similar to Brazil with the mean delay from symptom-onset-to-diagnosis decreasing overtime from 3.08 in epoch 1 and 2.62 in epoch 3 (Supplementary Table 2). However, there is substantially less variability in the delay from symptom-onset-to diagnosis and from hospitalisation-to-death (Figure 3 A and C). This can be seen by the mean difference in delay from symptom-onset-to diagnosis and from hospitalisation-to-death between the highest state (Nayarit) and lowest state (Chihuahua) differing only by 2.33 days and 3.76 days respectively over all epochs (Supplementary table 3). Further, like Brazil, we also found that increases in the mean delay from symptom-onset-to-diagnosis was negatively correlated with symptom-onset-to-death (r_s_ = -0.38, p = 0.03) and positively correlated with symptom-onset-to-hospitalisation (r_s_ = 0.65, p < 0.01) (Supplementary Figure 1).

#### Argentina

In contrast to both Brazil and Mexico, epoch 1 in Argentina had the lowest delay from symptom-onset-to-diagnosis and the highest delay for the symptom-onset-to-death (Supplementary Figure 2). We found that there was a high inter-state variance, as seen by the elongated shape on the violin plot. For the 11 states where data was available for the delay from symptom-onset-to-hospitalisation, the mean delay increased from 2.46 days in epoch 1 to 4.64 days in epoch 3 whilst the mean delay between symptom-onset-to-death decreased from 16.98 days in epoch 1 to 15.54 days in epoch 3 (Supplementary table 2). We did not find a significant relationship between delay distributions but note that no data was available for hospitalisation-to-death (Supplementary Figure 1).

#### Colombia

Like Argentina, we find that for Colombia epoch 1 had the lowest delay from symptom-onset-to-diagnosis (Supplementary Figure 3A). We found that the overall mean delay between symptom-onset-to-diagnosis is substantially longer for epoch 3 (10.83 days) than for epoch 1 (1.96 days) (Supplementary table 2). This large increase in the overall mean delay is driven by three states; Norte de Santander, Guainía, and Santa, which have mean delay from symptom-onset-to-diagnosis of over 30 days for epoch 3 (Supplementary Figure 3A, Supplementary Table 3). There is no overall trend across symptom-onset-to-death (Figure 5B).

#### Deconvolution of case time series

We apply methods from Goldstein et al. to raw SARS-CoV-2 case counts (date of confirmation) in the four countries studied to obtain the deconvolved daily case counts. Figure 4 shows the deconvolved incidences curves. Notability, we find a marked delay in cases for Colombia in epoch 3 particularly after the 1st of February 2021. Further, we find that the initial peak in cases within Brazil had significant delays perhaps due to high case incidence.

**Figure 4:**
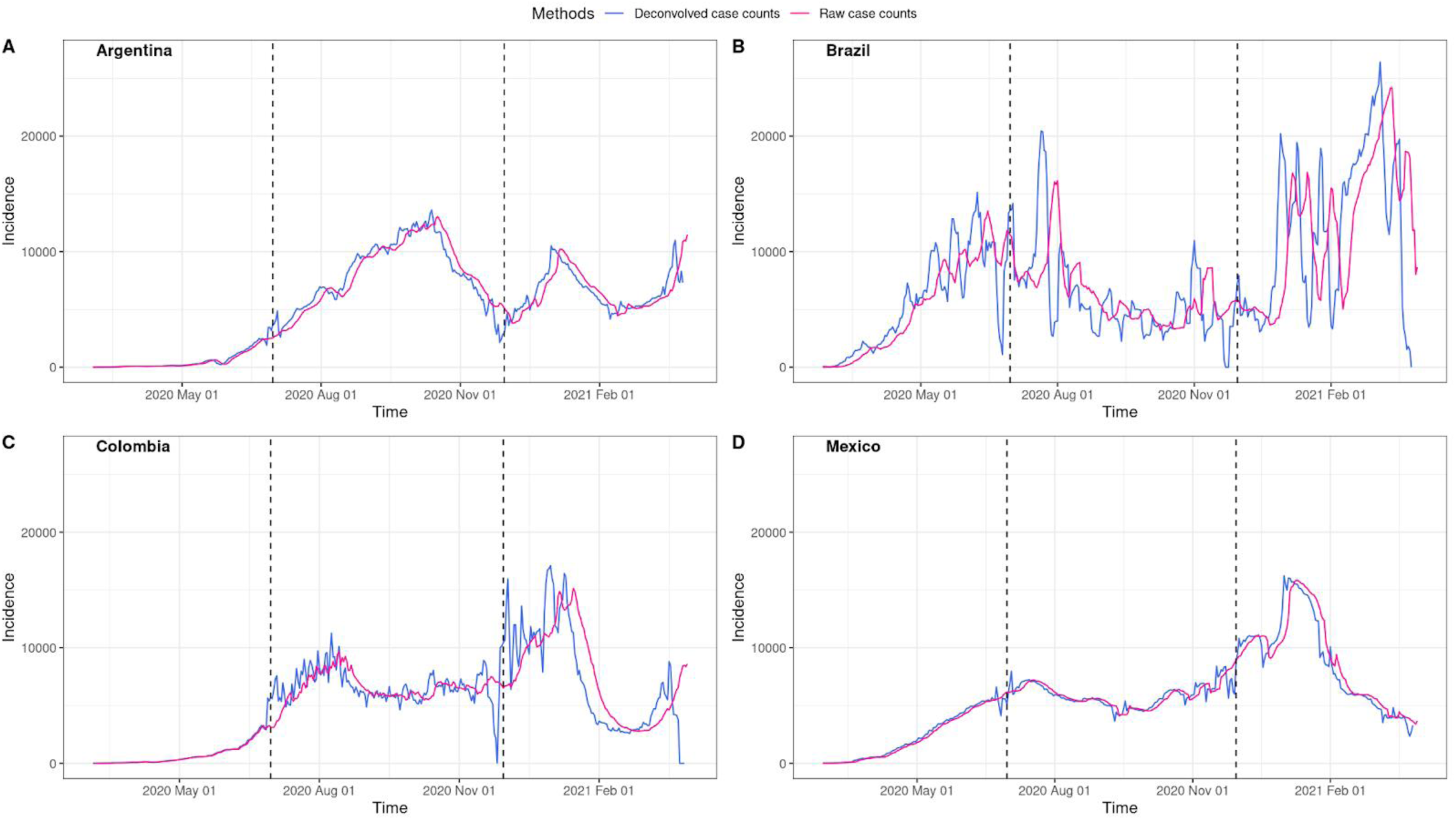
Deconvolved case counts have been estimated from raw case counts extracted from Global.health line lists for Argentina, Brazil, Colombia, and Mexico. The blue and red line represents a 7-day rolling average of deconvolved and raw case counts respectively. The dashed lines represent epoch change times.

#### Growth rates

We applied the Pellis *et al*. model to estimate *r*_*t*_ from raw case data and deconvolved case data for each of our countries of interest (Figure 5). Based on the deconvolved case counts, initially, for all countries the mean *r*_*t*_ was above zero, indicating a growing epidemic. For all countries the mean *r*_*t*_ declined moving into the second epoch. Argentina experienced a mean *r*_*t*_ falling consistently below zero during epoch 2. Towards the end of epoch 2, the mean *r*_*t*_ increased above zero and remained above zero at the start of epoch 3 for all countries.

**Figure 5:**
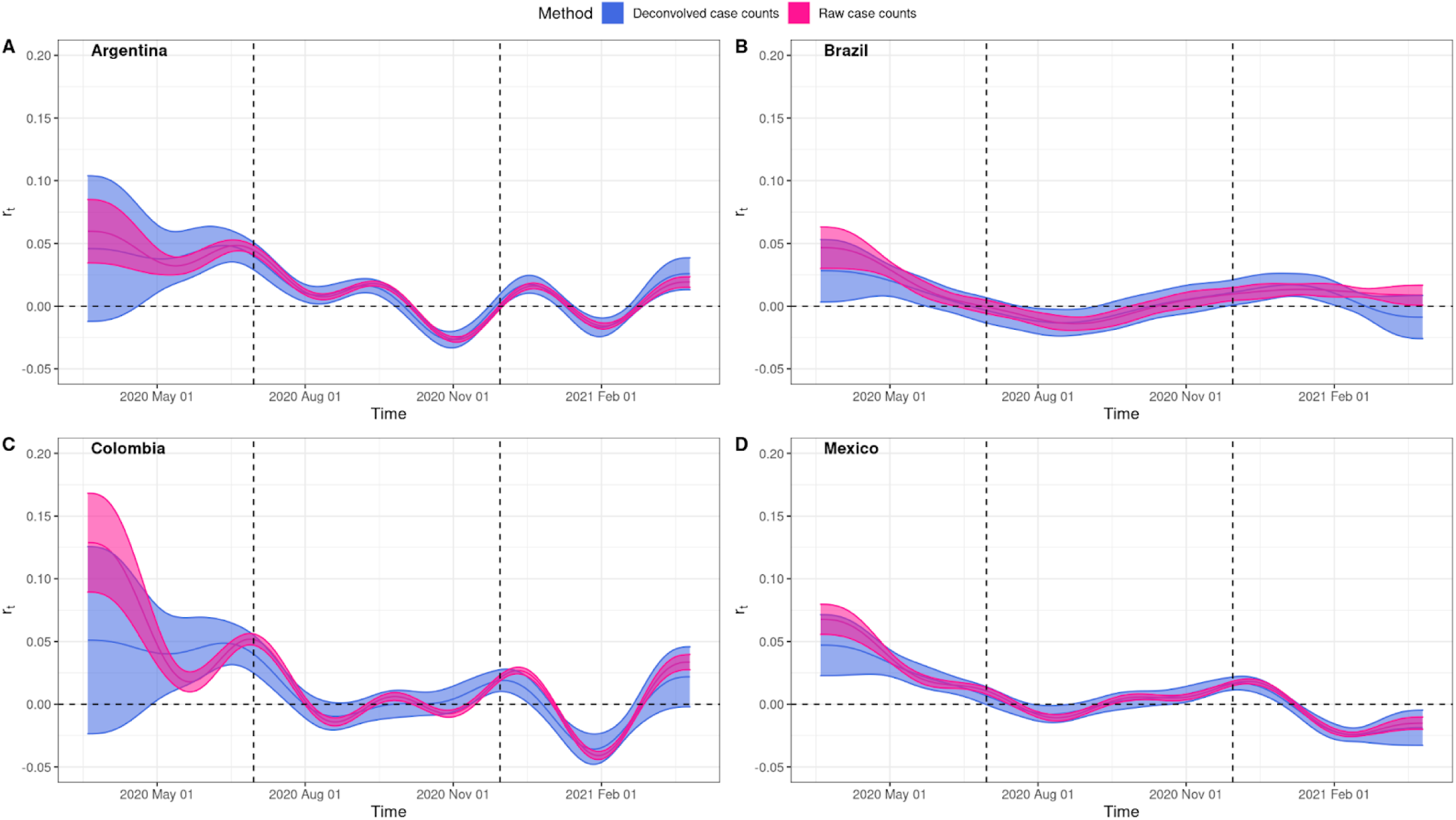
r_t_ estimated from both raw and deconvolved case counts for Argentina, Brazil, Colombia, and Mexico. The light-shaded area represents the 95% Confidence Interval with the darker-shaded area presenting where the two estimations overlap. The solid line represents the mean r_t_ estimate with r_t_ estimated from raw case counts in red and deconvolved case counts in blue. The vertical dashed lines represent epoch change times and the horizontal dashed line represents r_t_= 0.

Generally, it appears that the *r*_*t*_ estimated from the raw case counts lags behind the *r*_*t*_ estimated from the deconvolved case counts, which is expected. However, this difference is not significant, and all 95% confidence intervals (CIs) are overlapping (Figure 5). At the start of the study period there is an increase in uncertainty for the deconvolved case counts represented by the wider CIs and in general higher *r*_*t*_ in all countries using raw case data.

Next, we evaluated *r*_*t*_ on a state level by selecting states with the lowest mean delay (Figure 6A, 6B, 6E and 6G) and highest mean delay (Figure 6B, 6D, 6F and 6H) of symptom-onset-to-confirmation. We compared *r*_*t*_ estimates from state and national deconvolved case counts in addition to raw case counts. When the delay from symptom-onset-to-confirmation is low, there is a mismatch between the *r*_*t*_ calculated using national level deconvolved case counts and the *r*_*t*_ calculated using raw case and state level deconvolved case counts. For example, in La Pampa, Argentina (Figure 6E), mean *r*_*t*_ is initially below 0 (−0.03 d^-1^) when using national level deconvolved case counts and above 0 when using raw (0.1 d^-1^) and state level deconvolved case counts (0.07 d^-1^). Conversely, when the delay from symptom-onset-to-confirmation is high, there is a mismatch between the *r*_*t*_ calculated using state level deconvolved case counts and the *r*_*t*_ calculated using raw case and national level deconvolved case counts. This can be seen in Roraima state, Brazil (Figure 6B), where there are fluctuations of *r*_*t*_ below and above 0 when *r*_*t*_ is calculated using state level deconvolved case counts when compared to *r*_*t*_ estimations from raw and national level deconvolved case counts where *r*_*t*_ = ∼0 indicating epidemic stabilisation has occurred.

**Figure 6:**
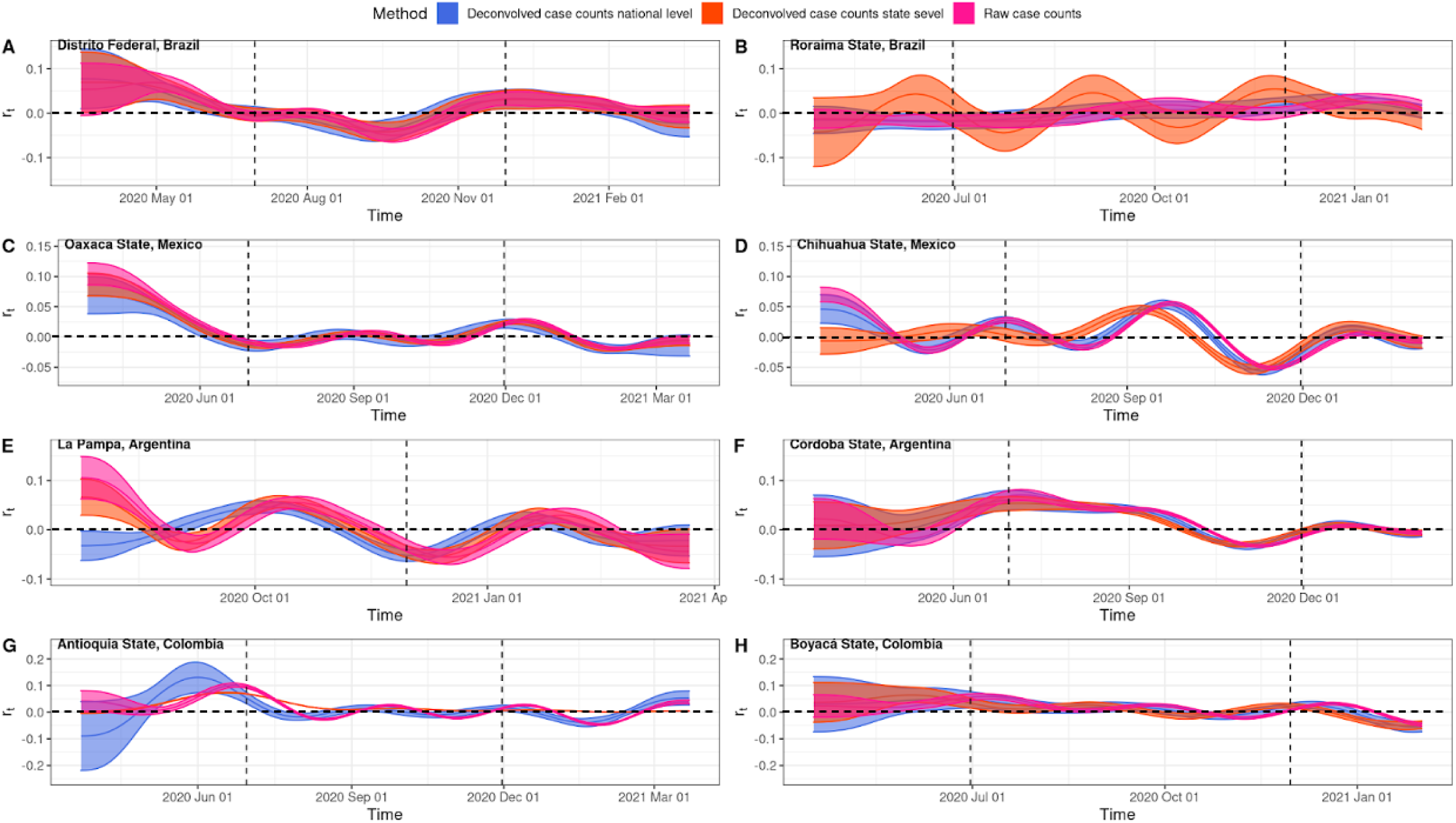
r_t_ estimated from both raw, national and states level deconvolved case counts for states with the highest mean delay in symptom-onset-to-diagnosis (6A,6C,6E and 6G) and the lowest mean delay in symptom-onset-to-diagnosis (6B,6D,6F and 6H) for Argentina, Brazil, Colombia, and Mexico. The light-shaded area represents the 95% Confidence Interval with the darker-shaded area presenting where the two estimations overlap. The solid line represents the mean r_t_ estimate with r_t_ estimated from raw case counts in red, state level deconvolved case counts in orange and national level case counts in blue. The vertical dashed lines represent epoch change times.

## Discussion

In this study, we fitted multiple probability density functions to a number of epidemiological datasets to quantify the delay from symptom-onset-to-hospitalisation and hospitalisation-to-death, from the Global.health database (https://global.health/), using Bayesian hierarchical models. Subsequently, the national level and state level delay from symptom-onset-to-confirmation was used to deconvolve raw case counts and we measure the impact on case growth rates *r*_*t*._

We found that across all countries investigated (Argentina, Brazil, Colombia, and Mexico) there were strong geographical heterogeneities between states for our inferred delays (Supplementary Table 2 and 3) with the delays from symptom-onset-to-diagnosis and symptom-onset-to-death being most accentuated. Whilst studies exploring testing heterogeneities in Latin America are limited, in the early stages of the epidemic, frequent and free testing was not available and testing was largely reserved for patients within hospitals and symptomatic individuals (Asahi, Undurraga and Wagner, 2021; Gaudart *et al*., 2021; Vandenberg *et al*., 2021). Less urbanised states, such as Roraima state, Brazil, Michoacán state, Mexico, and Boyacá, Colombia within the countries analysed had the largest delay in symptom-onset-to-diagnosis. It has been shown in other settings that access to testing varied geographically based on geographic accessibility (Jaitman, 2015) and length of travel to healthcare facilities (Syed, Gerber and Sharp, 2013; Kelly *et al*., 2016; Rader *et al*., 2020).

In addition to spatial heterogeneities, strong temporal heterogeneities were observed. For Brazil and Mexico, the delay in symptom-onset-to-diagnosis decreased over time by 23% and 15% respectively whilst for Argentina and Colombia this delay increased over time by 18% and 452% respectively. Brazil and Mexico experienced a more rapid epidemic progression with the first wave of cases peaking at the end of the first epoch (Figure 4B and 4D). In contrast, Colombia and Argentina had a slower epidemic progression with their first wave of cases peaking in the second epoch (Figure 4A and 4C). This is also reflected in the number of data entries with Brazil and Mexico having over double the number of entries in epoch 1 than Argentina and Colombia (Figure 1). With limited testing resources available (Asahi, Undurraga and Wagner, 2021; Gaudart *et al*., 2021; Vandenberg *et al*., 2021), it is plausible that public health departments in Brazil and Mexico struggled to test all symptomatic cases in a timely manner when compared to Argentina and Colombia which had fewer cases during that period.

By using deconvolution to infer the unlagged time series of infections, we can improve the accuracy of key epidemiological parameters (Gostic *et al*., 2020). In particular, by using the delay distribution of symptom-onset-to-confirmation we allow *r*_*t*_ to be estimated closer to real time (some have called this ‘nowcasting’ (McGough *et al*., 2020)). We found that in states with a small delay from symptom-onset-to-diagnosis there was a mismatch between *r*_*t*_ estimated using national level deconvolved case counts and raw and state level deconvolved case counts. Further, in states with a large delay from symptom-onset-to-diagnosis there was a mismatch between *r*_*t*_ estimated using state level deconvolved case counts and raw and national level deconvolved case counts. This is significant as using deconvolved case counts at a less granular spatial scale can significantly affect the interpretation of the epidemic picture. For example, for Roraima state, Brazil (Figure 6B) using national level deconvolved case counts to estimate *r*_*t*_ we would predict that epidemic stabilisation has occurred even though cases have changed significantly throughout time (https://github.com/CSSEGISandData/COVID-19).

While our results provide a rigorous underpinning and insight into delay distributions and impact of these on epidemiological parameters estimation, we acknowledge several limitations. The Global.health database which contains line lists that our distributions have been estimated from, though extensive, contains typing errors, and the degree to which these bias our estimates are unknown. Our data ingestion pipeline is mostly automated and only occasionally are we able to manually verify the accuracy of the data. Further, when comparing line list data between and within countries we note disparities in notification systems and differences in case definitions. Further work should evaluate the demographic biases in these data and how that may affect transmission dynamics (longer delays for less severe cases in younger age groups may impact transmission substantially). Lastly, there is a low testing rate for the countries analysed (Hasell et al., 2020) and heterogeneities in testing rates in both time and space (Vandenberg *et al*., 2021) which can influence the results for both cases and *r*_*t*_. Future epidemiological work is needed to compare parameters estimated from case data, death data and excess death data across different settings (Gostic *et al*., 2020) and more intensive monitoring and/or the use of alternative data sources such as genomic data (Inward, Faria and Parag, 2022) is needed to improve the reliability of estimations. Few countries report highly detailed epidemiological data limiting the ability to perform robust analyses on the impact of delays on transmission across the world. One primary concern for limited sharing of these data is privacy. Our work demonstrates the ability to perform scalable analyses of delay distributions and their impact on case growth rates and could be applied across all settings and through time. In the future, raw data may not need to be shared publicly: algorithms could locally process line list data stored in each country, with only aggregated statistics shared globally.

This work has highlighted the impact that both spatial and temporal heterogeneities can have on delay distributions and subsequent estimations of the case growth rate. Whilst more epidemiological datasets from a variety of countries and regions with different sampling intensities are needed to create a more generalisable understanding and to identify predictors of these differences, we have shown that accounting for delays on both a national and state level can introduce substantial differences in the estimation of epidemiological parameters. This finding identifies the need for more targeted attempts at performing epidemiological surveillance and epidemic analyses particularly in resource-poor settings which have limited surveillance systems.

## Data Availability

All data used within this study are open source with details of how to obtain them found in the methods section.

## Role of the Funding Sources

M.U.G.K. is supported by The Branco Weiss Fellowship - Society in Science, administered by the ETH Zurich and acknowledges funding from a Google Faculty Award, the Oxford Martin School. This work was partially funded by the European Union Horizon 2020 project MOOD (#874850), Google.org, and the Rockefeller Foundation. The contents of this publication are the sole responsibility of the authors and do not necessarily reflect the views of the European Commission.

## CRediT authorship contribution statement

M.U.G.K, R.P.D.I and F.J conceived and designed the study, R.P.D.I and F.J performed the analyses. G.L., A.L.B., A.D. and F.J. assisted with data curation, ingestion, and processing. R.P.D.I and F.J wrote the manuscript which was edited and supervised by M.U.G.K. All authors have contributed to and approved the manuscript for submission.

## Conflicts of interest

The authors declare no conflicts of interest.

## Supplementary Information

**Supplementary Table 1:**
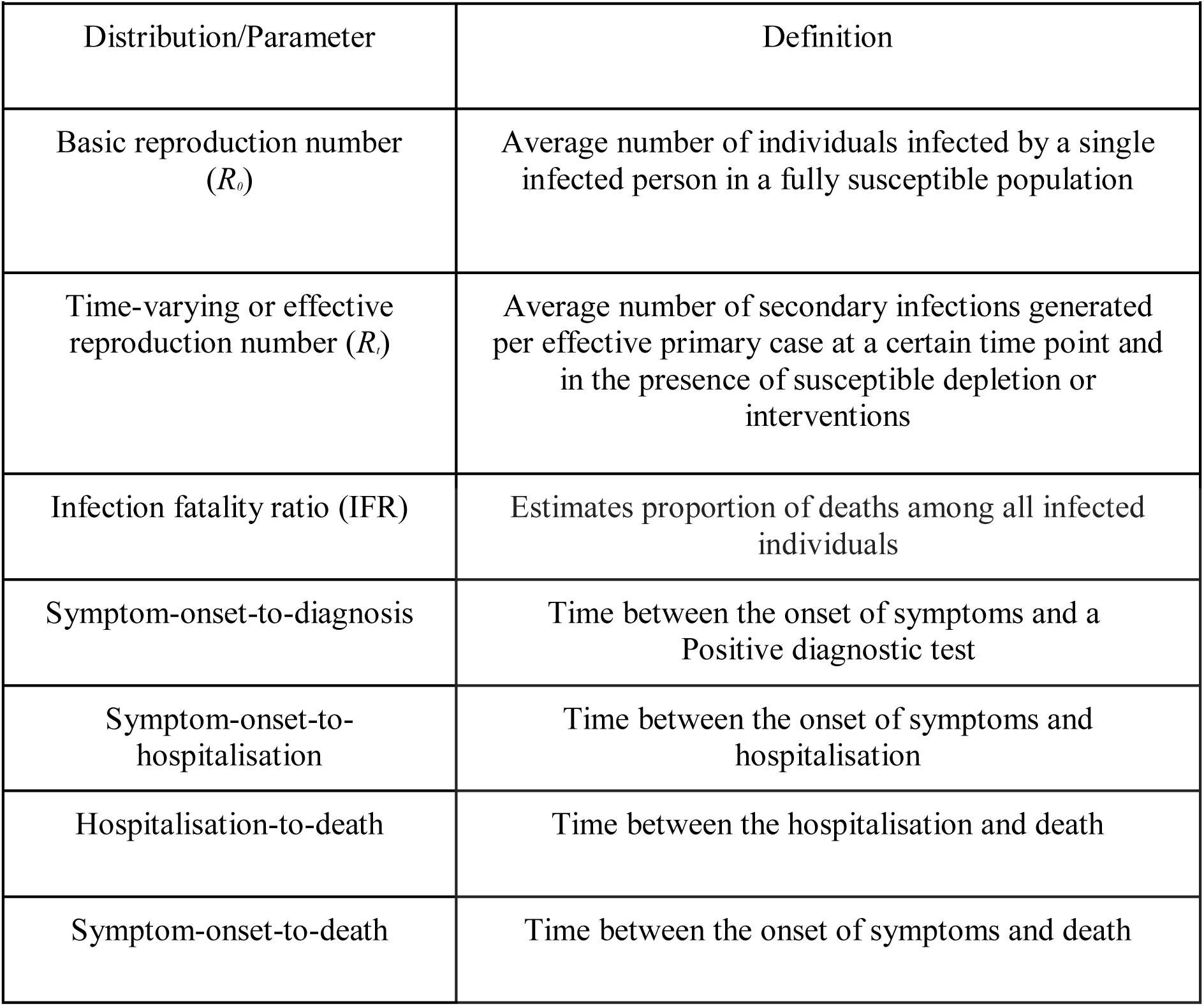
Key distribution, parameters, and definitions for SARS-CoV-2

**Supplementary Table 2:** Summary of inferred means and case counts per country and epoch

[Impact of spatiotemporal heterogeneity in COVID-19 disease surveillance on epidemiological parameters and case growth rates](https://docs.google.com/spreadsheets/d/1fg8yKO19cCxW-rWjPmksz9ydSV2gO44jWwI-cKkAcKM/edit#gid=1916821368)

**Supplementary Table 3:** Summary of inferred means at the state level per country and epoch for each distribution of interest

[Impact of spatiotemporal heterogeneity in COVID-19 disease surveillance on epidemiological parameters and case growth rates](https://docs.google.com/spreadsheets/d/1fg8yKO19cCxW-rWjPmksz9ydSV2gO44jWwI-cKkAcKM/edit#gid=1916821368)

**Supplementary Figure 1:**
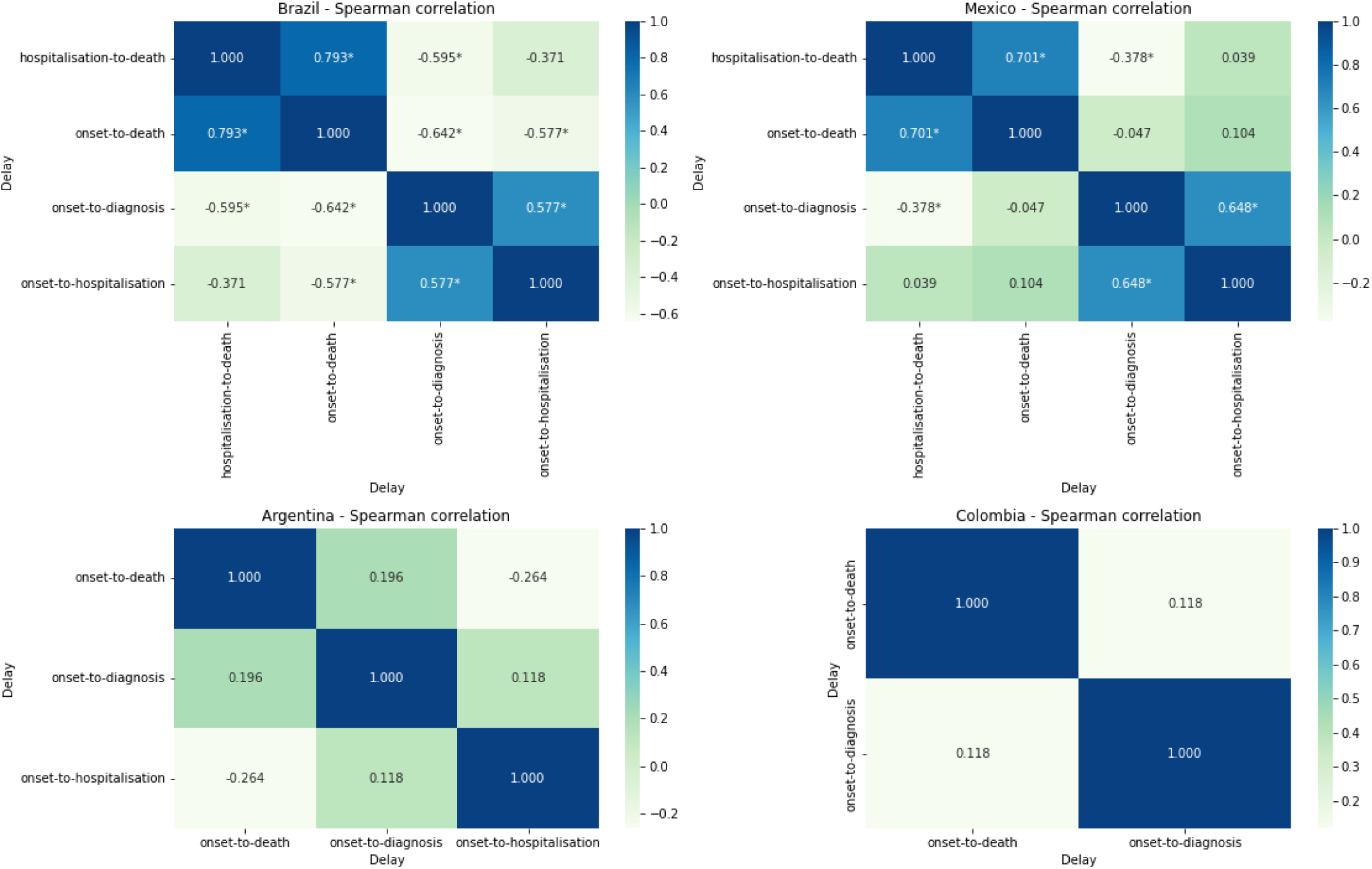
Spearman’s rank-order correlation coefficient correlations between delay distributions for each state, taking the mean value of the delay across all epochs. Values marked with a star denote p-value of this pairwise correlation was less than 0.05.

**Supplementary Figure 2:**
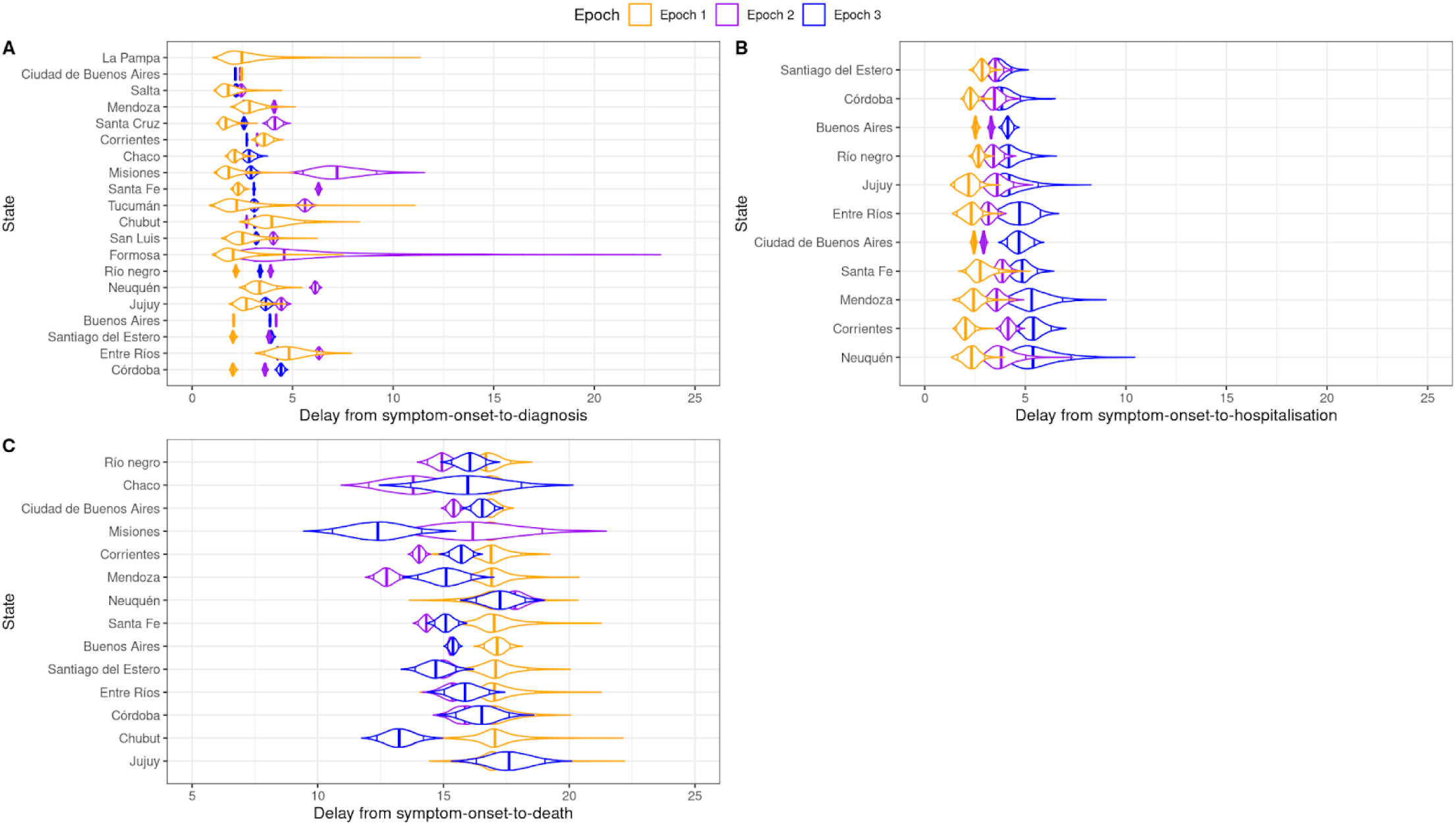
Delay distributions are estimated from daily case counts on the state level for three distinct epochs for Argentina. Supplementary Figure 2A, 2B and 2C represent the delay from symptom-onset-to-diagnosis, -hospitalisation, and -death respectively. Orange represents epoch 1, purple represents epoch 2 and blue represents epoch three. All plots are ordered from the smallest to largest by the epoch with the smallest mean delay.

**Supplementary Figure 3:**
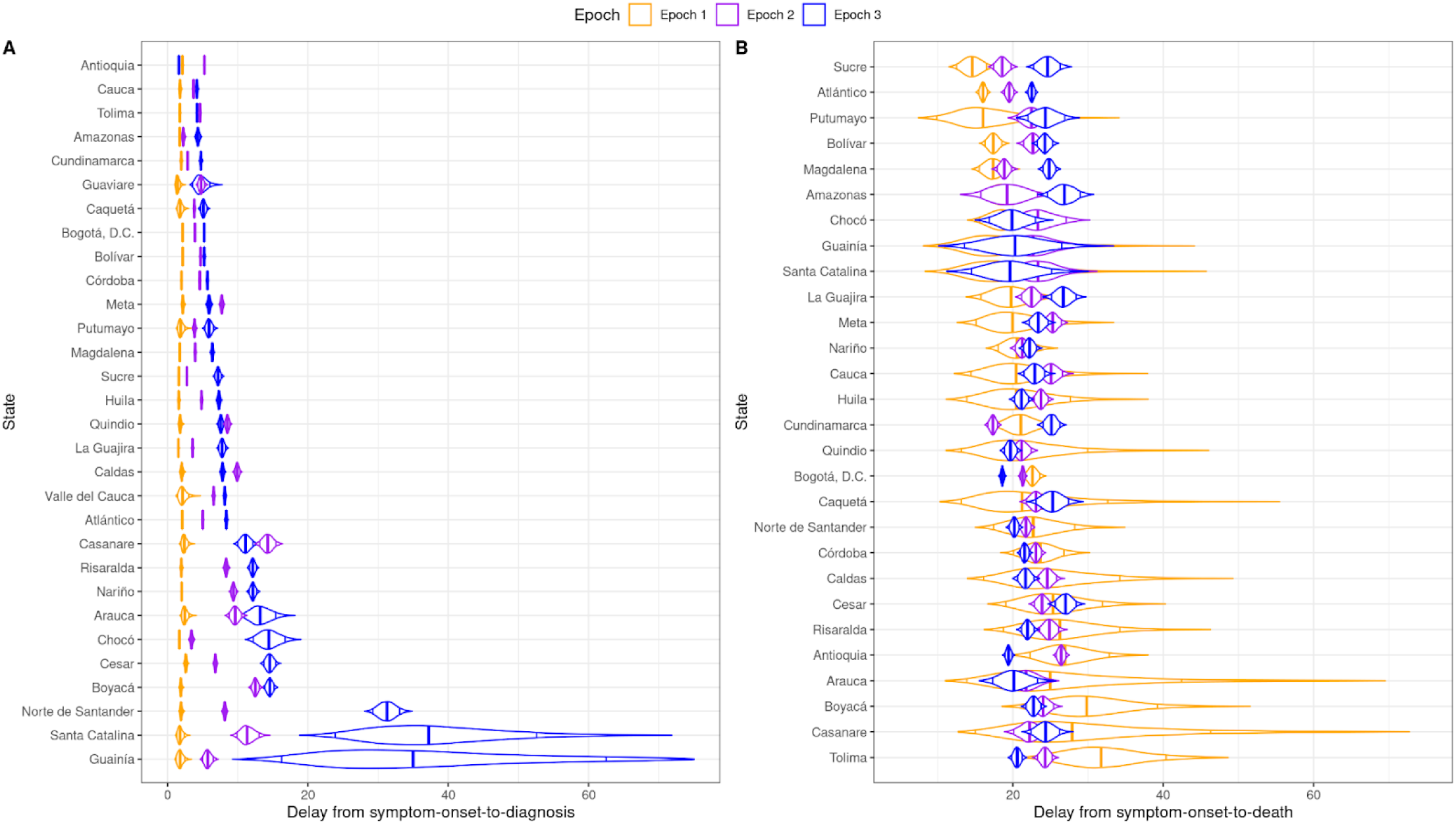
Delay distributions are estimated from daily case counts on the state level for three distinct epochs for Colombia. Supplementary Figure 3A and 3B represent the delay from symptom-onset-to-diagnosis and -death respectively. Orange represents epoch 1, purple represents epoch 2 and blue represents epoch three. All plots are ordered from the smallest to largest by the epoch with the smallest mean delay.

## Notes

### Competing Interest Statement

The authors have declared no competing interest.

### Summary of Updates

Ordering of authors updated

